# Antemortem vs. postmortem histopathological and ultrastructural findings in paired transbronchial biopsies and lung autopsy samples from three patients with confirmed SARS-CoV-2 infection

**DOI:** 10.1101/2020.12.30.20248929

**Authors:** D Gagiannis, VG Umathum, W Bloch, C Rother, M Stahl, HM Witte, S Djudjaj, P Boor, K Steinestel

**Affiliations:** Department of Pulmonology, Bundeswehrkrankenhaus Ulm, 89081 Ulm, Oberer Eselsberg 40, Germany; Institute of Pathology and Molecular Pathology, Bundeswehrkrankenhaus Ulm, 89081 Ulm, Oberer Eselsberg 40, Germany; Department of Molecular and Cellular Sport Medicine, German Sport University Cologne, 50933 Cologne, Am Sportpark Müngersdorf 6, Germany; Department of Hematology and Oncology, Bundeswehrkrankenhaus Ulm, 89081 Ulm, Oberer Eselsberg 40, Germany; Department of Hematology and Oncology, University Hospital Schleswig-Holstein Campus Luebeck, 23552 Luebeck, Ratzeburger Allee 160, Germany; Institute of Pathology, RWTH Aachen University Hospital, 52074 Aachen, Pauwelsstrasse 30, Germany

**Author notes:** **Corresponding author: Konrad Steinestel, MD PhD**, Institute of Pathology and Molecular Pathology, Bundeswehrkrankenhaus Ulm, Oberer Eselsberg 40, 89081 Ulm, Germany. **Shared first authorship** (D Gagiannis, VG Umathum). **Declaration of interest:** DG and KS were speakers for Boehringer-Ingelheim. All other authors declare no conflict of interest.

**Keywords:** COVID-19, SARS-CoV-2, ARDS, lung pathology, diffuse alveolar damage, organizing pneumonia, lung fibrosis

## Abstract

**Background:** Acute respiratory distress syndrome (ARDS) is the major cause of death in coronavirus disease 2019 (COVID-19). Multiple autopsy-based reports of COVID-19 lung pathology describe diffuse alveolar damage (DAD), organizing pneumonia (OP) and fibrotic change, but data on early pathological changes as well as during progression of the disease are rare.

**Research question:** Comparison of histopathological and ultrastructural findings in paired transbronchial biopsies (TBBs) and autopsy material from three patients with confirmed SARS-CoV-2-infection.

**Methods:** We prospectively enrolled 3 patients with confirmed SARS-CoV-2 infection. Full clinical evaluation was performed including high-resolution computed tomography (HR-CT). We took TBBs at different time points during the disease and autopsy tissue samples after the patients’ death.

**Results:** SARS-CoV-2 was detected by RT-PCR and/or FISH in all TBBs. Lung histology revealed pneumocyte hyperplasia and capillary congestion in one patient who died short after hospital admission with detectable virus in 1/2 autopsy samples from the lung. SARS-CoV-2 was detected in 2/2 autopsy samples from a patient with a fulminant course of the disease and very short latency between biopsy and autopsy, both showing widespread DAD. In a third patient with a prolonged course, i.e. five weeks of ICU treatment with ECMO, autopsy samples showed extensive interstitial fibrosis without detectable virus by RT-PCR and/or FISH.

**Interpretation:** We report the course of COVID-19 in paired TBB and autopsy samples from three patients at an early stage, in rapidly progressive and in a prolonged disease course. Our findings illustrate vascular, organizing and fibrotic patterns of COVID-19-induced lung injury and suggest an early spread of SARS-CoV-2 from the upper airways to the lung periphery with diminishing viral load during disease.

---

Since SARS-CoV-2 and the resulting disease, COVID-19, emerged in late 2019, much effort has been put in a better understanding of the clinical course of the disease. While most patients present with a rather mild symptoms, some patients – especially those sharing risk factors such as older age, cardiovascular disease or chronic obstructive pulmonary disease (COPD) – are at risk of developing life-threatening respiratory failure ^1^. Early autopsy studies conducted in China described diffuse alveolar damage (DAD) with an early edematous phase followed by hyaline membrane formation, desquamation of pneumocytes and an increased interstitial mononuclear infiltrate ^2^. In one case, Tian et al. report loose intra-alveolar fibromyxoid proliferation reminiscent of organizing pneumonia (OP). In the meantime, it is widely accepted that COVID-19 follows a biphasic pattern of an initial viral response phase followed by an inflammatory second phase, and that mortality is linked primarily to the development of acute respiratory distress syndrome (ARDS) ^14^. Based on a meta-analysis of 131 reported autopsy cases, Polak et al. postulated that the main histologic patterns of COVID-19-related lung injury can be categorized into epithelial (reactive changes and DAD), vascular (microvascular damage, thrombi and OP) and fibrotic ^5^, however these patterns may overlap and be coexistent in the same patient at a given time point. Nicholson et al. proposed that an initial (pre-)exudative phase of DAD (0-7 days) is followed by an organizing phase (1 week to months) and might ultimately progress to fibrosis (months) ^6^. The exact mechanisms of SARS-CoV-2-related ARDS development are not fully understood. Is has been postulated that severity of COVID-19 might correlate with a hyper-inflammatory response and uncontrolled secretion of cytokines, showing similarities to cytokine releasing syndrome (CRS) ^7^; however, it has been shown that cytokine levels in severe cases of COVID-19 are lower compared to severe influenza patients ^8^. Our own group demonstrated overlapping clinical, serological, and imaging features between severe COVID-19 and lung manifestation of autoimmune disease such as systemic lupus erythematosus (SLE) or systemic sclerosis ^9^. In our study, the presence of autoantibodies (antinuclear antibodies and extractable nuclear antibodies, ANA/ENA) was significantly associated with a need for intensive care treatment and the occurrence of severe complications. This finding has now been confirmed by others and might be attributed to extrafollicular B cell activation with excessive production of antibody-secreting cells (ASCs) in critically ill COVID-19 patients ^10-12^. A better understanding of the pathophysiology of lung injury in COVID-19 would also shed light on the urgent question of long-term sequelae of the disease in millions of COVID-19 survivors. Despite a large number of autopsy studies which represent a snapshot of the disease at the time of death, to the best of our knowledge, there is so far no study comparing *antemortem* vs. *postmortem* histopathological and ultrastructural features of COVID-19. We report here the histopathology of transbronchial biopsies and autopsy material together with RT-PCR- and FISH-based detection of SARS-CoV-2 and ultrastructural analyses from three patients with confirmed SARS-CoV-2 infection.

## METHODS

We consecutively included three (n=3) patients with positive SARS-CoV-2-RT-PCR (mucosal swab) admitted to the Bundeswehrkrankenhaus (German Armed Forces Hospital) Ulm in March and April 2020 after obtaining informed consent. Patients or their relatives had given written informed consent to routine diagnostic procedures (serology, bronchoscopy, radiology) as well as (partial) autopsy in the case of death, respectively, as well as to the scientific use of data and tissue samples in the present study. This project was approved by the local ethics committee of the University of Ulm (ref. no. 129-20) and conducted in accordance with the Declaration of Helsinki.

### Clinical characteristics

We collected clinical information from electronic patient files. Data included disease-related events, preexisting comorbidities, imaging, and clinical follow-up. Baseline clinical characteristics are given in **Table 1**. The “Berlin definition’’ was used to categorize ARDS.^13^ The Horovitz quotient (PaO_2_/FiO_2_) was assessed in all ARDS cases based on arterial blood gas analysis. During ICU treatment, ventilation parameters, duration of invasive ventilation, catecholamine support, prone positioning, Murray lung injury score and the need for additional temporary dialysis were continuously assessed^14^. A profitable trial of prone positioning was defined by an increasing Horovitz quotient of 30 mmHg or more. One entire trial covered 16 hours of sustained prone positioning.

**Table 1.**
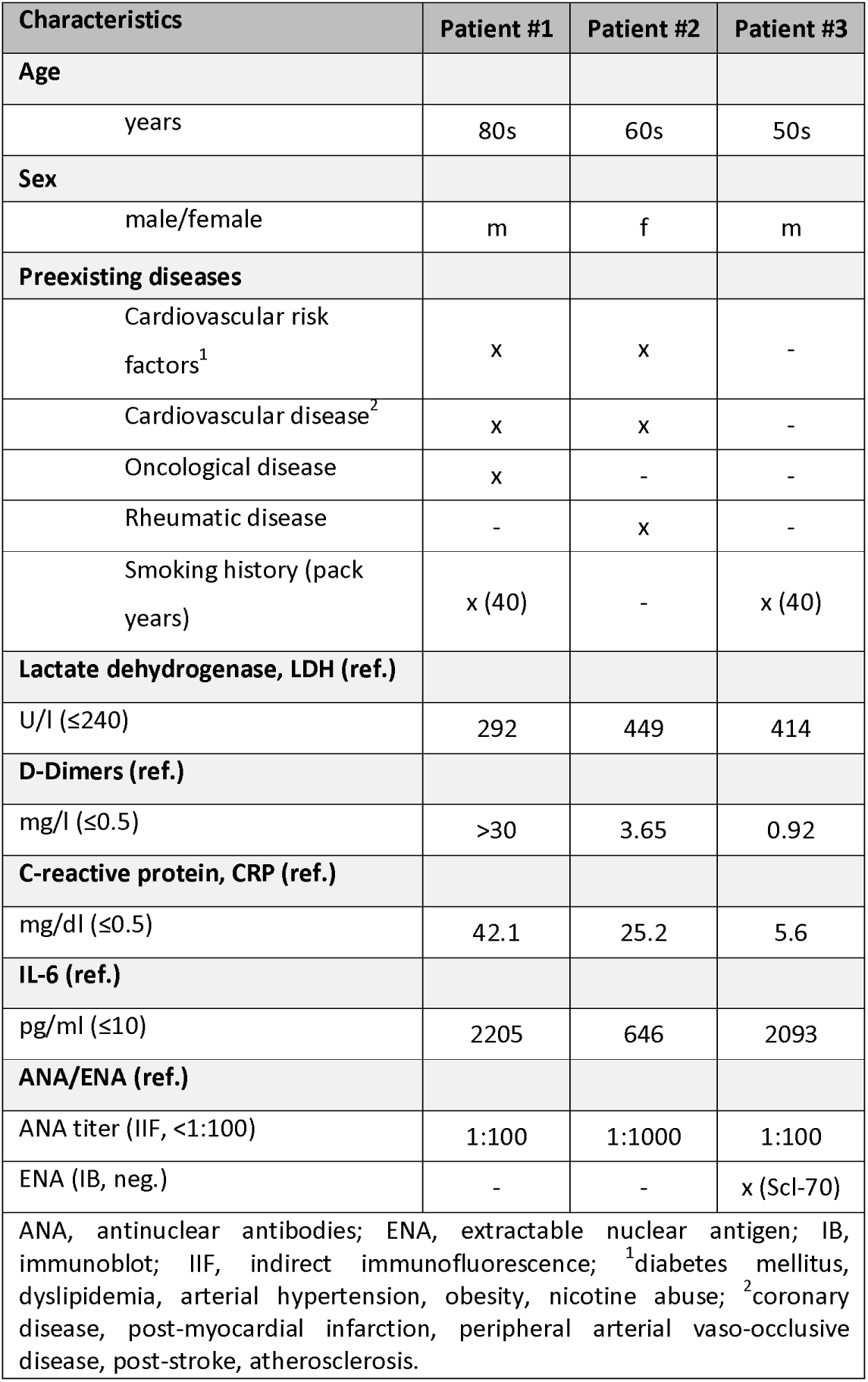
Clinical characteristics of COVID-19 patients.

### Serology/Laboratory values

Laboratory values upon admission to ICU included D-dimers, lactate dehydrogenase (LDH), interleukin-6 (IL-6) and C-reactive protein (CRP) (**Table 1**). ANA/ANCA/ENA screening was performed as previously described^9^.

### Imaging

Imaging was performed on a Somatom Force Scanner (Dual Source Scanner 2*192 slices, Siemens, Erlangen, Germany) in accordance with the guidelines of the German Radiological Society and our hospital’s COVID-19 guidelines, using low-dose CT (computed tomography) with high-pitch technology.^15^ The following parameters were used: Tube voltage: 100kV with tin filtering, tube current: 96 mAs with tube current modulation. In two cases examination was performed as a non-contrast enhanced full-dose protocol because of suspected ILD, in one case as a contrast-enhanced CT scan to exclude pulmonary thromboembolism. X-ray examinations were performed at the respective wards as bed-side X-ray examinations (Mobilett Mira Max, Siemens, Erlangen, Germany) as a single anterior-posterior view. The CT images were evaluated according to the Expert Consensus Statement of the RSNA and classified as typical, indeterminate, atypical and negative appearance for COVID- 19.^15 16^

### Histology and SARS-CoV-2 detection

Lung tissue specimens were obtained as transbronchial biopsies. In three deceased patients, partial autopsies were performed in which lung (central and peripheral areas), heart and liver tissue were sampled extensively. Specimens were stained with haematoxylin-eosin (HE), Phosphotungstic-Acid-Hematoxylin (PTAH), Elastica- van-Gieson (EvG) and Masson-Goldner (MG). For SARS-Cov-2 fluorescence in situ hybridization (FISH) Paraformaldehyde-fixed, paraffin-embedded 1µm sections of transbronchial biopsies (TBBs) and lung autopsy material were deparaffinized followed by dehydration with 100% ethanol. FISH was performed with the RNAscope^®^ Multiplex Fluorescent Reagent Kit v2 assay (Advanced Cell Diagnostics, Inc.) according to the maufacturer’s instruction. Briefly, a heat-induced target retrieval step followed by protease was performed. Afterwards sections were incubated with the following RNAscope^®^ Probe -V-nCoV2019-S (#848561- C1), -V-nCoV2019-S-sense (#845701-C1), -Hs-ACE2-C2 (#848151-C2) and -Hs-TMPRSS2-C2 (#470341-C2). After the amplifier steps the fluorophores OpalTM 570 and 650 (PerkinElmer Life and Analytical Sciences) were applied to the tissues incubated with C1 and C2 probe, respectively. Finally, nuclei were stained with DAPI and the slides were mounted with ProLongTM Gold antifade reagent (Invitrogen). Section were analyzed with Zeiss Axio Imager 2 and image analysis software (ZEN 3.0 blue edition). SARS-CoV-2 RNA was extracted using Maxwell^®^ 16 FFPE Plus Tissue LEV DNA Purification KIT (Promega) on Maxwell^®^ 16 IVD Instrument (Promega). Using TaqMan^®^ 2019-nCoV kit (ThermoFisher), detection of SARS-CoV-2 E-gene was performed according to the manufacturer’s instructions.

### Electron microscopy

Lung tissue was immersion-fixed with 4% paraformaldehyde in 0.1M PBS, pH 7.4. After several washing steps in 0.1M PBS, tissue was osmicated with 1% OsO4 in 0.1 M cacodylate and dehydrated in increasing ethanol concentrations. Epon infiltration and flat embedding were performed following standard procedures. Methylene blue was used to stain semithin sections of 0.5 µm. Seventy to ninety-nanometer-thick sections were cut with an Ultracut UCT ultramicrotome (Fa. Reichert) and stained with 1% aqueous uranylic acetate and lead citrate. Samples were studied with a Zeiss EM 109 electron microscope (Fa. Zeiss) coupled to a TRS USB (2048×2048, v.596.0/466.0) camera system with ImageSP ver.1.2.6.11 (x64) software

## RESULTS

### Baseline patient characteristics and clinical course of the disease

The study included two male and one female patient who were hospitalized for RT-PCR confirmed SARS-CoV-2 infection at the Bundeswehrkrankenhaus Ulm, Germany. The timeline of disease course is depicted in **Fig. 1 A**. Baseline laboratory values and clinical characteristics are summarized in **Table 1**. Of note, ANA screening was positive in all three patients; in patient #3, a specific autoantibody (Scl-70) could be detected by immunoblot. While patients #1 and #3 had to undergo invasive ventilation 2 and 3 days after the diagnosis of COVID-19, patient #2 was transferred from another hospital in a critical state. In patient #1, TBB was performed before knowledge of a positive SARS-CoV-2 test result (day 0). This patient died from pulmonary thromboembolism 5 days later. Except for the continuation of a treatment protocol with hydroxychloroquine and azithromycin that had been established in the previous hospital for patient #2, no specific therapeutic regimens were administered to patients #1 and #3. TBBs in patient #2 and #3 were performed after 13 and 23 days of ICU treatment, respectively, to assess the fibrotic change of lung parenchyma and to evaluate a possible use for antifibrotic treatment options. An Autopsy was performed in all three cases on the day after the patients’ death following the published guidelines^17^.

**Figure 1.**
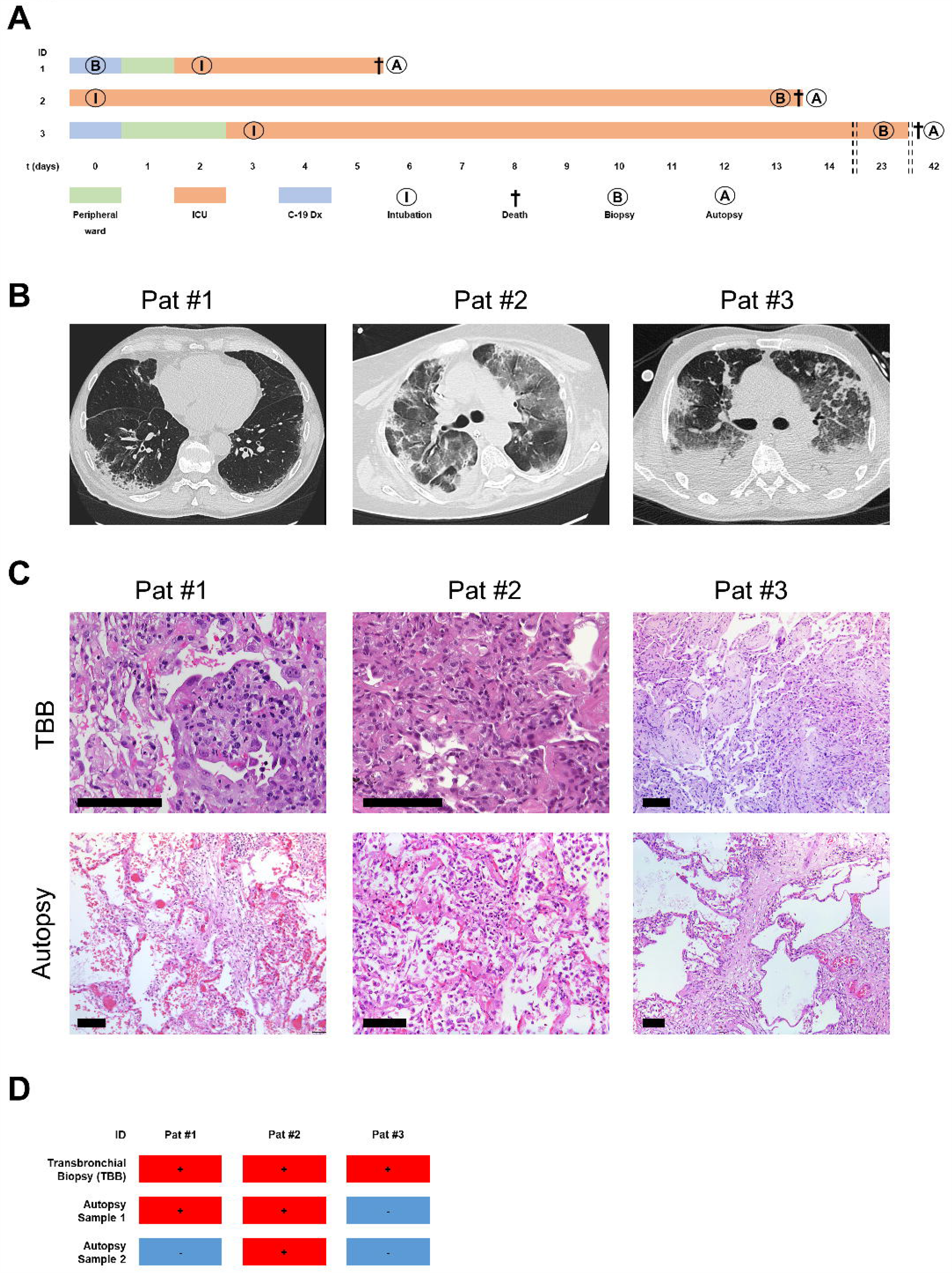
A, Timeline of the disease course in three COVID-19 patients. While patients #1 and #3 were treated in the peripheral ward (green) after diagnosis (blue), patient #2 was taken over from another hospital while immediate intubation (I) was required. Periods in the intensive care unit (ICU) ranged from four to 39 days (orange). Transbronchial biopsies (B) were taken on the day of hospital admission (patient 1), day 14 (patient 2) and day 23 (patient 3). Autopsies (A) were performed one day after death in all 3 patients. **B, Results from imaging**. Imaging in patient# 1 showed an NSIP-like pattern with minimal ground-glass-opacities (GGOs) in the left subpleural space. In patient# 2, there were diffuse GGOs and both subpleural and peribronchial consolidations with a positive aerobronchogram resembling organizing pneumonia. Imaging in patient #3 showed parenchymal lung injury with diffuse GGOs, bronchiectasis, cysts, and air-trapping. **C, Representative histology and CT imaging**. TBBs in patient #1 showed reactive pneumocyte changes (“Napoleon hat sign”) and loose fibromyxoid plugging with capillary congestion in autopsy samples; in patient #2, TBBs showed alveolar collapse with fibrin deposition and plug-like fibromyxoid organization (AFOP-like pattern) in autopsy samples. TBBs from patient #3 showed condensed fibromyxoid plugging of alveoli, while autopsy samples show interstitial collagen deposition and pseudo-honeycombing. *Scale bar in all images*: *100µm*. **D, Results from SARS-CoV-2 testing by RT-PCR**. While transbronchial biopsies (TBB) were positive in all patients, 1/2 and 2/2 lung autopsy samples were positive in patients #1 and #2, respectively. All autopsy samples were SARS-CoV-2-negative in patient #3.

### Imaging

Representative CT scans from all patients are shown in **Fig. 1 B**. While there was a discrete NSIP-like pattern with only minimal ground-glass opacities (GGO) in patient #1, patient #2 showed far more widespread GGOs combined with consolidations and a positive aerobronchogram, suggestive of OP. In patient #3, imaging showed severe parenchymal damage with diffuse GGOs, bronchiectasis, cyst formation and air trapping.

### Histopathologic findings in vital transbronchial biopsies (TBB) and lung autopsy samples

The main finding in TBBs from patient #1 was reactive changes of pneumocytes (multinucleated cells, “Napoleon hat sign”) together with a discrete interstitial mononuclear infiltrate. In some alveoli, there was accumulation of fibrin without hyaline membrane formation (**Fig. 1 C**). Autopsy samples from the same patient showed focal capillary congestion together with microthrombosis and very few fibromyxoid plugs in the alveolar lumen. TBBs from patient #2 - who was already mechanically ventilated at the time of biopsy - showed alveolar collapse with entrapment of fibrin as well as reactive changes in few pneumocytes. Autopsy samples from the same patient showed widespread fibromyxoid plugging with entrapment of ball-like fibrin and only very few residual ventilated alveoli. In some alveoli, there was a shedding of reactive pneumocytes. In patient #3, TBBs showed extensive OP with only very sparse interstitial inflammation. There was extensive interstitial fibrosis and pseudo-honeycombing in autopsy samples from patient #3 with the formation of subepithelial fibroblast foci.

### SARS-CoV-2 testing on tissue samples and SARS-CoV-2/ACE2/TMPRSS2-FISH

RT-PCR analyses from tissue samples detected SARS-CoV-2 in TBBs from all three patients and in both autopsy samples from patient #2(**Fig. 1 D**). SARS-CoV-2 testing from autopsy samples revealed positivity in one out of two samples in patient #1, while both autopsy samples were negative in patient #3. In line with that, SARS-CoV- 2 could be detected by FISH in TBBs, but not in autopsy samples from patients #1 and #3 (**Fig. 2 A-B** and **E-F**). The virus was detected in airway epithelial cells but not in pneumocytes in patient #1. SARS-CoV-2 was detected in both TBBs and autopsy samples from patient #2 (**Fig. 2 C-D**). There was only a weak and focal signal for angiotensin converting enzyme-2 (ACE2) in all investigated samples (**Fig. 2 A’’-F’’**), while the signal for transmembrane protease serine subtype 2 (TMPRSS2) was strongly detectable in all TBBs and autopsy samples from patient #2, correlating with the presence of SARS-CoV-2 (**Fig. 2 A’’’-F’’’**)

**Figure 2.**
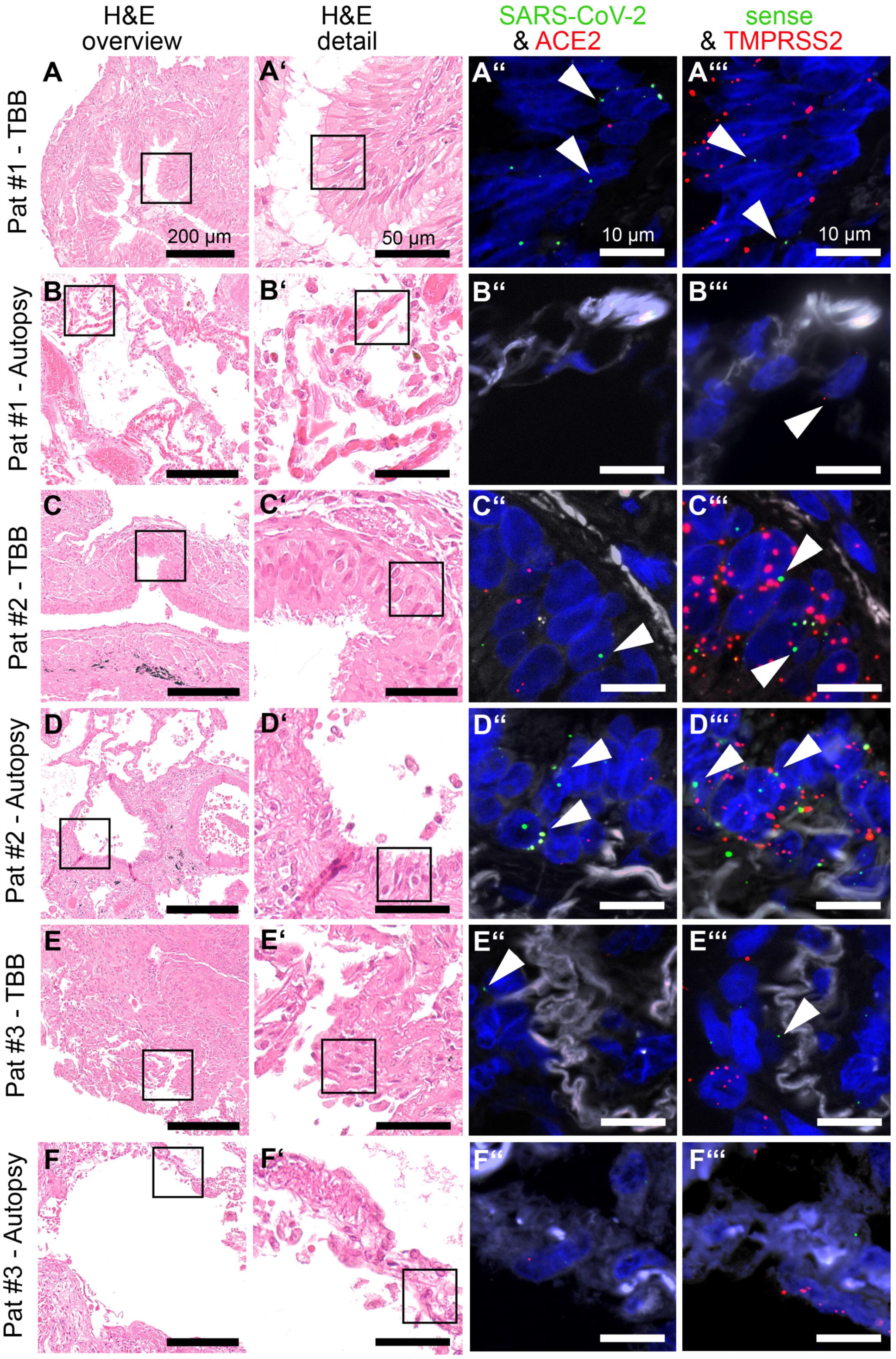
Fluorescence in situ hybridization (FISH) of SARS-Cov-2. H&E staining and two combinations of FISH that target SARS-CoV-2 S gene+ Angiotensin-converting enzyme 2 (ACE2) and replicative SARS-CoV-2-S gene sense + Transmembrane protease, serine 2 (TMPRSS2) have been performed on consecutive slides. SARS-CoV-2 (green, A’’-F’’) is strongly expressed in the respiratory epithelium as shown in the biopsy of patient #1 (arrows in A’’) and 2 (arrows in C’’) and the autopsy of patient #2 (arrows in D’’). To a lower extent virus could be detected in alveolar epithelial cells as shown in the biopsy of patient #3 (arrow E’’). High viral replication is detectable in patient #2 as visualized with the V-nCoV2019-S sense probe (green, C’’’ and D’’’). The receptor TMPRSS2 is strongly expressed, mainly in the respiratory epithelium similar to the virus (red, A’’’-F’’’) much stronger than ACE2 (red, third A’’-F’’), whereas ACE2 is only weakly expressed.

### Ultrastructural findings in lung autopsy samples

Electron microscopy (EM) was performed on autopsy samples from all three patients. There was capillary congestion with erythrocytes and fragmentocytes in patient #1. Of note, we observed long and thin cytoplasmic protrusions of erythrocytes consistent with acanthocytosis. There was only minimal collagen deposition in the interstitium (**Fig. 3 A** and **B**). In patient #2, there was widespread desquamation of alveolar epithelium, interstitial edema and deposition of loosely organized collagen in the interstitium (**Fig. 3 C**). There was extensive extracellular matrix deposition containing collagen and elastic fibrils in patient #3 (**Fig. 3 D**). Moreover, we found luminal extension of endothelial protrusions consistent with intussusceptive (splitting) angiogenesis (**Fig. 3 E**).

**Figure 3.**
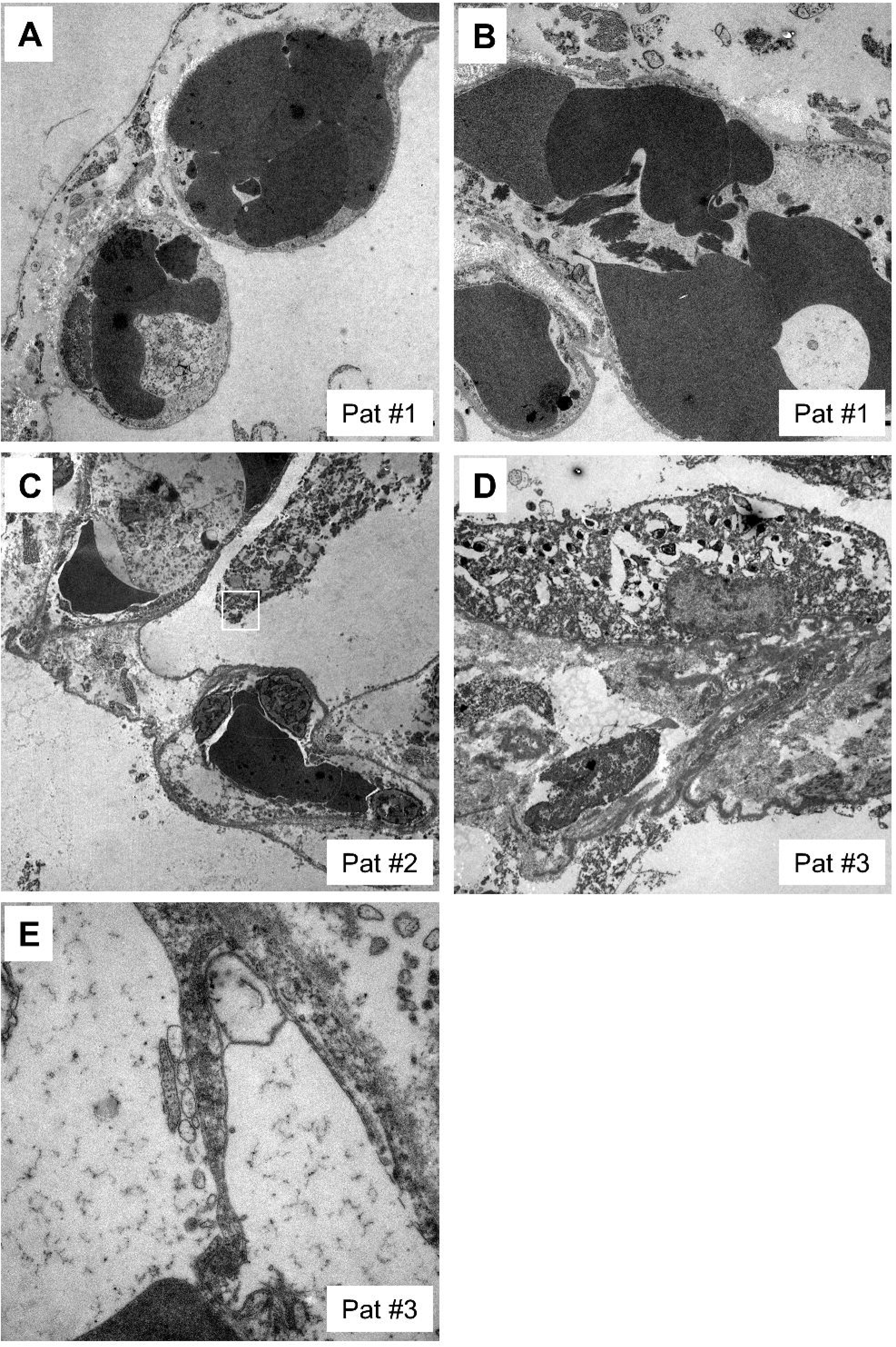
Ultrastructural analyses. **A**, Alveolar capillaries are filled with erythrocytes and fragmentocytes (asterisks) embedded in an alveolar septum with slightly increased extracellular matrix accumulation. *Original magnification: 4000x, image width: 23,5µm*. **B**, Capillaries are filled with erythrocytes and fragmentocytes (asterisks). The erythrocytes are altered in shape and form very long and thin cytoplasmic processes as a hint of a special kind of acanthocytosis (arrows). *Original magnification: 3000x, image width: 31,5µm*. **C**, Alveolar septum with erythrocytes and leucocyte-filled capillaries is enlarged by interstitial fluid accumulation and loosely organized fibrillar (arrows) and non-fibrillar extracellular matrix (arrowheads). The septum is bounded by alveolar basement membrane with sheared off alveolar epithelium. *Original magnification: 3000x, image width: 31,5µm*. **D**, Interstitial fibrosis of a thickened alveolar septum with fibroblast debris (asterisks) and an adjacent alveolar epithelial cell (A). The extracellular matrix contains collagen fibrils, elastin and an accumulation of basement membrane material. *Original magnification: 3000x, image width: 31,5µm*. **E**, The alveolar capillary endothelium reveals cytoplasmic processes with the formation of an interendothelial contact (arrow). The cytoplasmic process divides the lumen as a sign of intussusceptive (splitting) angiogenesis. *Original magnification: 20000x, image width: 5,9µm*.

## DISCUSSION

In the current paper, we report on antemortem and postmortem pathology in three patients with confirmed SARS-CoV-2 infection including severe courses of COVID-19. From a clinical viewpoint, all three patients shared established risk factors for a severe disease course (middle or advanced age, cardiovascular risk factors/disease and/or history of smoking)^18^. Laboratory findings at the point of ICU admission showed elevated levels for LDH, D-dimers, IL-6, and CRP, in line with published data^19^. Of note, two of the three patients (patients #2 and #3) were positive for autoantibodies (ANA titers ≥1:320 and/or positive ENA immunoblot). Previously, our group showed that detection of autoantibodies is associated with a need for intensive care (ICU) treatment and the occurrence of severe complications in COVID-19^9^. Since all patients were treated in the spring of 2020, no immunosuppressive agents (dexamethasone) had been administered.

Histopathologic findings in lung biopsies from living COVID-19 patients have rarely been reported. A very early report described oedema, proteinaceous exudate, focal reactive hyperplasia of pneumocytes with patchy inflammation, and multinucleated giant cells in two patients who underwent lobectomy for lung cancer and who were retrospectively found to have COVID-19 at the time of operation^3^. We observed similar changes in patient #1 who was also not yet known to have COVID-19 at the time of biopsy, indicating that pneumocyte hyperplasia with proteinaceous exudate represents the earliest response to SARS-CoV-2 infection. The virus was detected by RT-PCR and FISH while the latter method demonstrated SARS-CoV-2 in bronchiolar epithelium where it was co-expressed with TMPRSS2, but not ACE2. This is in line with data from the literature showing that TMPRSS2-expressing cells are highly susceptible to SARS-CoV-2 infection and TMPRSS2 expression might correlate with age, sex and smoking habits^20 21^. We observed low ACE2 expression in airway epithelial cells in patient #1, consistent with previous reports^22^.

Only 1 of 2 autopsy samples from patient #1 tested positive for SARS-CoV-2 in RT-PCR, and characteristic histopathological changes were very focal, supporting the assumption that the patient died early during the disease before widespread involvement of the lung. Such intrapulmonary heterogeneity of SARS-CoV-2 infection has previously been described^23^. Since the cause of death was pulmonary thromboembolism with high D-dimer levels (>30 mg/l, ref. ≤0.5 mg/l), it is very interesting that electron microscopy showed capillary congestion with erythrocytes and fragmentocytes in this patient. Endothelial inflammation and activation of coagulation are associated with multiorgan failure and increased mortality, and antithrombotic drugs have been proposed as potential therapies to prevent thrombosis in COVID-19^24^. Given these findings, we think that patient #1 might represent the vascular pattern of COVID-19-associated lung injury which has been shown to occur early in the course of the disease^5^ and that these patients might profit from anti-coagulative therapy at an early time point.

While patient #1 represents an early phase of the host response to SARS-CoV-2 infection, both TBBs and autopsy samples from patient #2 showed widespread DAD with ball-like-fibrin and fibromyxoid plugging. This acute fibrinous and organizing pneumonia (AFOP)-like pattern has previously been reported in response to SARS-CoV-2 infection, and is believed to represent an intermediate form of lung injury between exudative DAD and OP^6 25^. In line with that, we think that the pattern of lung injury we observed in this patient represents the climax of lung injury in response to SARS-CoV-2 infection, and the virus could be detected in all samples from this patient by RT-PCR and FISH, respectively. TMPRSS2, but not ACE2, was highly expressed in both respiratory epithelium and pneumocytes in TBBs and autopsy samples. It has to be noted that the uniformity of the histologic findings might reflect the short time span between TBB and autopsy (48h) in this patient. Ultrastructural analyses revealed the deposition of collagen fibrils in the interstitium, predicting the development of interstitial fibrosis. While we know now that the application of corticosteroids would be beneficial for patients with SARS-CoV-2 induced organizing lung damage, it is unclear whether such treatment would also prevent the development of fibrosis. In patient #3, TBBs on day 23 showed extensive organizing pneumonia and only sparse inflammation, while autopsy samples after the patient’s death on day 42 showed extensive interstitial fibrosis and microscopic honeycombing. The virus was still detectable in TBBs, but no virus could be detected by RT-PCR and FISH in autopsy samples. Ultrastructural analyses confirmed extensive collagen deposition. These findings support previous reports describing long-term follow-up of severe COVID- 19 and underline the potential of the development of interstitial fibrosis^26^. Interestingly, in our previous study, 5/6 specific autoantibodies in patients with severe COVID-19 were associated with some form of sclerosing

CTD, raising the question if a pro-fibrotic (auto)immune response contributes to the development of lung fibrosis in COVID-19 patients^9^. EM further confirmed the presence of intravascular endothelial protrusions consistent with intussusceptive (splitting) angiogenesis as reported before^27^.

## INTERPRETATION

Taken together, we show here antemortem and postmortem biopsies from three patients with severe COVID- 19, illustrating histopathological changes during the disease (**Fig. 4**). Early changes include pneumocyte hyperplasia and capillary congestion, which may be a sign of the risk of thromboembolic complications, especially when high D-dimers are present. At this point, the virus has not necessarily spread throughout the lung. The climax of lung injury is widespread DAD with or without organization, possibly including intermediate AFOP-like patterns; at this point, virus is present in the airways and the lung. When this phase is survived, there is increasing honeycomb-like fibrosis, however, viral load decreases over time and SARS-CoV-2 may not be detected in late-stage disease. Development of lung fibrosis as a long-term consequence of COVID-19 is associated with high morbidity and mortality and should definitely be circumvented whenever possible, raising the question of a possible use of anti-fibrotic therapies, such as pirfenidone, in survivors of severe COVID-19.

**Figure 4.**
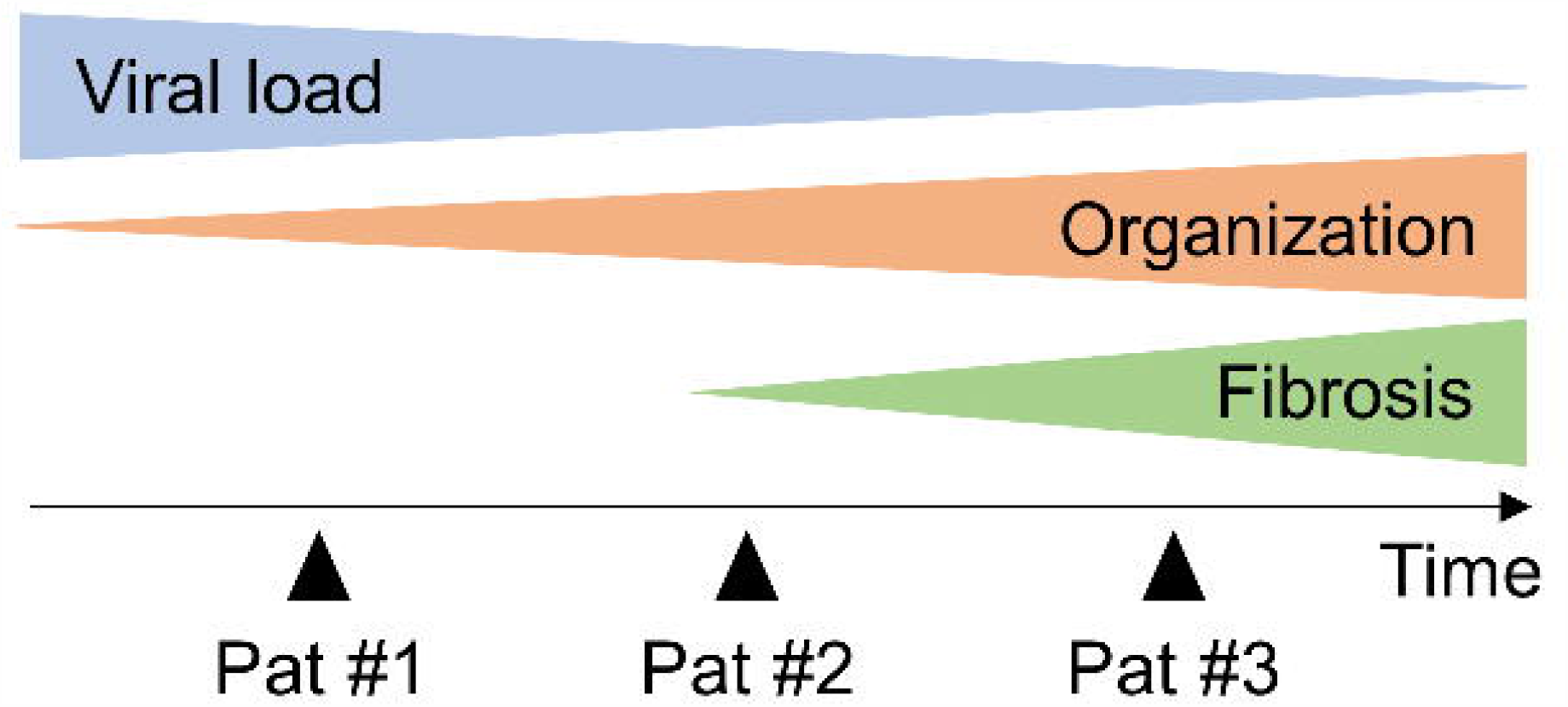
Schematic model for the course of COVID-19 and possible implications for long-term sequelae in the lung. While SARS-CoV-2 viral load decreases over time, an increasing and dysregulated (auto-)inflammatory response exerts deleterious effects on lung function and implies the risk for development of progressive interstitial fibrosis. The different stages in disease progression are reflected by the observed changes in patients 1-3.

## Data Availability

All relevant data included in the manuscript. Original slides/tissue samples are archived at the senior author's institution.

## ACKNOWLEDGEMENTS

The authors would like to thank all patients and their families for their consent to the use of data and images in the present study. We further thank Carsten Hackenbroch, MD, for providing imaging data. The authors are grateful for the outstanding quality of care of COVID-19 patients provided by the team of the intensive care unit (ICU) at the Bundeswehrkrankenhaus Ulm.

## AUTHOR CONTRIBUTIONS

Study concept: DG, VGU and KS. Data collection: DG, VGU, WB, CR, MS, HMW, SD, PB, KS. Sample collection: DG, VGU, WB. Initial draft of manuscript: KS. Critical revision and approval of final version: all authors.

